# Humoral Immunogenicity of Three COVID-19 mRNA Vaccine Doses in Patients with Inflammatory Bowel Disease

**DOI:** 10.1101/2021.12.22.21268217

**Authors:** Trevor L. Schell, Keith L. Knutson, Sumona Saha, Arnold Wald, Hiep S Phan, Mazen Almasry, Kelly Chun, Ian Grimes, Megan Lutz, Mary S. Hayney, Francis A. Farraye, Freddy Caldera

**Affiliations:** Department of Medicine, Division of Gastroenterology and Hepatology, University of Wisconsin School of Medicine & Public Health, Madison, Wisconsin, United States; Department of Immunology, Mayo Clinic Jacksonville, Florida, United States; Department of Medicine, University of Wisconsin School of Medicine & Public Health, Madison, Wisconsin, United States; LabCorp, R&D and Specialty Medicine; School of Pharmacy, University of Wisconsin-Madison, Madison, Wisconsin, United States; Inflammatory Bowel Disease Center, Division of Gastroenterology and Hepatology, Mayo Clinic, Jacksonville, Florida, United States

**Author notes:** **Corresponding author:** Freddy Caldera, DO, MS, 1685 Highland Avenue, Madison, WI, USA 53705-2281. **Guarantor of the article:** Freddy Caldera, DO, MS. **Specific author contributions:** T.L.S. – acquisition of data, analysis and interpretation of data, drafting of the manuscript, and critical revision of the manuscript. F.C. – study concept and design, acquisition of data, analysis and interpretation of data, drafting of the manuscript, and critical revision of manuscript. K.L.K – analysis and interpretation of data and critical revision of manuscript. S.S. – critical revision of manuscript. A.W. – critical revision of manuscript. H.S.P – acquisition of data, critical revision of manuscript. M.A. – acquisition of data, critical revision of manuscript. K.C. – acquisition of data, critical revision of manuscript, I.G. – critical revision of manuscript. M.L. – critical revision of manuscript. M.S.H – study concept and design, acquisition of data, analysis, and interpretation of data, drafting of the manuscript, and critical revision of manuscript. F.A.F. – analysis and interpretation of data and critical revision of manuscript.

**Keywords:** Inflammatory bowel disease, Crohn’s disease, Ulcerative colitis, COVID-19 vaccine

## Abstract

Herein, we evaluated the humoral immunogenicity of a third COVID-19 mRNA vaccine dose in patients with IBD. All patients were seropositive and had higher antibody concentrations after the third dose than after completion of the two-dose primary series.

## Introduction

Three safe and effective COVID-19 vaccines were authorized by the Food and Drug Administration in response to the COVID-19 pandemic. Neither patients with inflammatory bowel disease (IBD) nor immunosuppressed patients were included in the original Phase III clinical trials. Studies have since demonstrated a humoral immune response rate of 95–99% following vaccination with a two-dose mRNA vaccine series in patients with IBD.^1,2^ Results show that those on certain immune-modifying therapies, such as anti-tumor necrosis factor (anti-TNF) therapy in combination with an immunomodulator or the use of systemic corticosteroids, exhibit a relatively diminished humoral immune response and a relative decrease in serum antibody concentrations over time.^1–3^

For those who are moderately-to-severely immunocompromised, the Advisory Committee on Immunization Practice (ACIP) recommends a three-dose primary mRNA vaccine series. The aim of this study was to evaluate the humoral immunogenicity of a third COVID-19 mRNA vaccine dose in patients with IBD. We hypothesized that they would mount a significant humoral immune response, and that those on certain immune-modifying therapy, such as systemic corticosteroids or combination therapy, may have lower antibody concentrations.

## Methods

This was a multicenter, prospective, non-randomized study comprised of patients with IBD and healthy controls (HC) in the “<u>H</u>umo<u>R</u>al and <u>C</u>ell<u>UL</u>ar initial and <u>S</u>ustained immunogenicity in patients with IBD” (HERCULES) cohort.^2^ Participants with IBD were enrolled at the University of Wisconsin-Madison (Madison, Wisconsin) and Mayo Clinic (Jacksonville, Florida), while HC were employees of LabCorp. Patient eligibility criteria included a diagnosis of IBD, age 18–85 years, stable doses of maintenance therapy (≥ 2 months), completion of a two-dose mRNA vaccine series confirmed by interview, review of medical records, and if applicable, review of the Wisconsin Immunization Registry. HC eligibility criteria included absence of immunosuppressive therapy and documented completion of a two-dose mRNA vaccine series. A third COVID-19 mRNA vaccine dose was available to patients with IBD but not HC. No participants had clinical history of COVID-19 infection, and those with laboratory evidence of prior infection, as demonstrated by presence of SARS-CoV-2 nucleocapsid antibodies, were excluded.

The primary outcome was total serum SARS-CoV-2 anti-spike IgG antibody concentrations following a third dose compared to antibody concentrations following the two-dose series in the IBD cohort. Secondary outcomes included antibody concentrations following a third dose in patients with IBD compared to antibody concentrations 180 days after the two-dose series in HC. The effects of vaccine manufacturer and immunosuppressive therapy on antibody concentrations following a third dose in patients with IBD were also evaluated.

Specific antibodies measured in sera were nucleocapsid and spike protein S1 receptor-binding domain (RDB)-specific IgG antibodies reported as mcg/ml as previously described.^2^ In patients with IBD, we measured antibody concentrations 28–35 days (t_1_) after completion of the two-dose series and 28–65 days (t_2_) after the third dose. Only patients with IBD who received a third dose had antibody concentrations measured at t_2_, and not every subject who had antibody concentrations measured at t_2_ had them measured at t_1_ due to timing of enrollment. In HC, we measured antibody concentrations 30 days (t_1_) and 180 days (t_2_) after completion of the two-dose series.

IBD treatment groups were defined as subjects on stable doses of maintenance therapy as previously described.^2^ Non-immunosuppressive therapy was defined as absence of IBD therapy, mesalamine monotherapy, or vedolizumab monotherapy. Immunosuppressive therapy was defined as thiopurine monotherapy (i.e., azathioprine, 6-mercaptopurine), anti-TNF monotherapy, anti-TNF combination therapy (i.e., plus antimetabolite), ustekinumab monotherapy or combination therapy, tofacitinib, or systemic corticosteroid therapy (i.e., any of the aforementioned groups plus systemic corticosteroids). Antibody concentrations between groups were compared using Mann-Whitney U tests. The study received IRB approval at the University of Wisconsin, Mayo Clinic, and LabCorp.

## Results

One hundred thirty-nine patients with IBD completed the two-dose series and had antibody concentrations measured at t_1_, and 85 received a third dose and had antibody concentrations measured at t_2_ (Table 1). Forty-six HC completed the two-dose series and had antibody concentrations measured at both timepoints. In the IBD cohort, 48.2% and 51.8% received the two-dose Moderna and Pfizer series, respectively, compared to 93.5% and 6.5% in HC (p<0.001). Two patients with IBD switched from the Moderna two-dose series to the Pfizer third dose. The median age of patients with IBD was significantly lower than that of HC (median 38 (IQR 30–49) vs 42 (IQR 35–58), p=0.033). The age of patients with IBD who received a third dose was significantly greater than that of those who completed only the two-dose series (median 48 (IQR 38–60) vs 41 (IQR 34–52), p=0.003). The characteristics of the groups were otherwise similar.

**Table 1:**
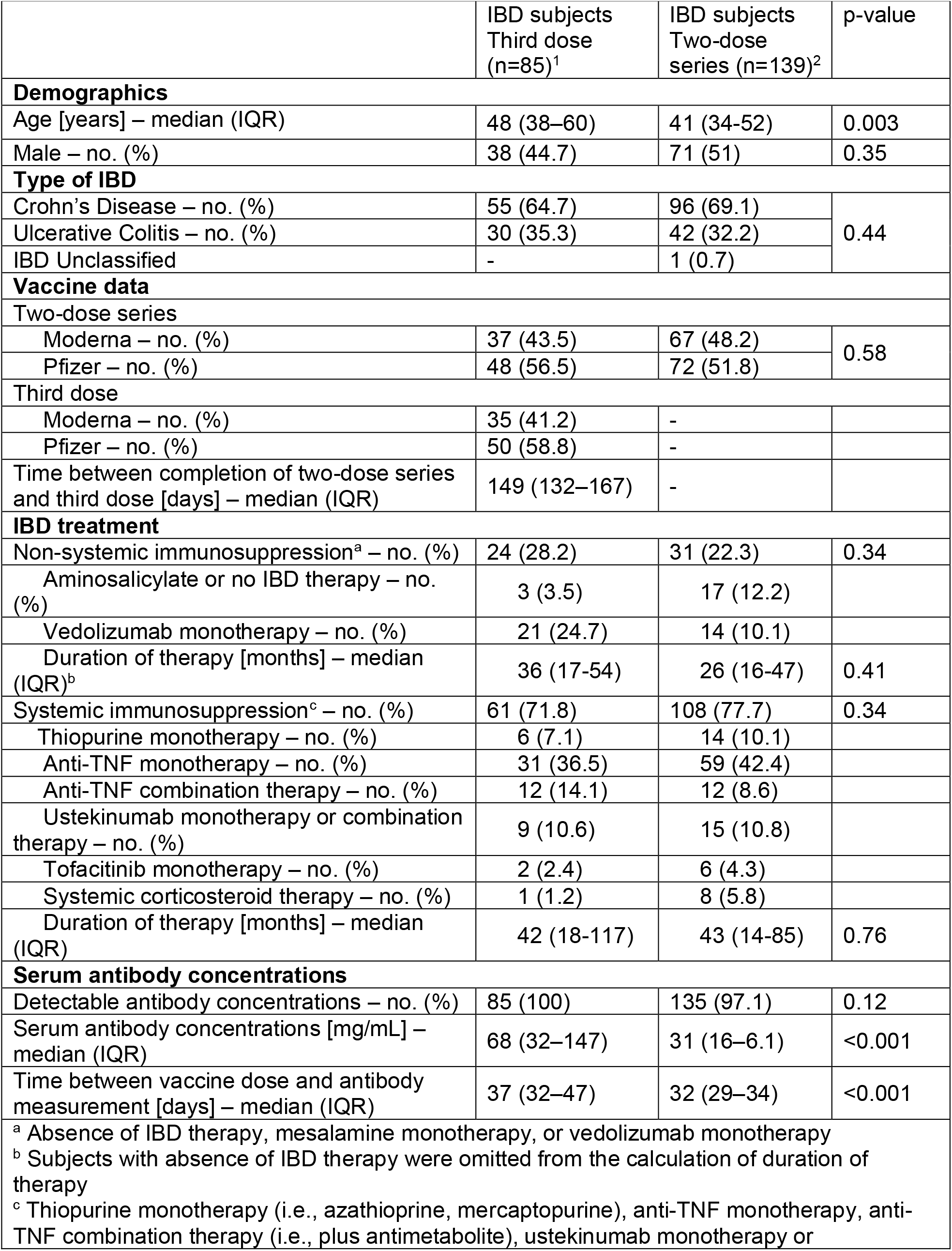
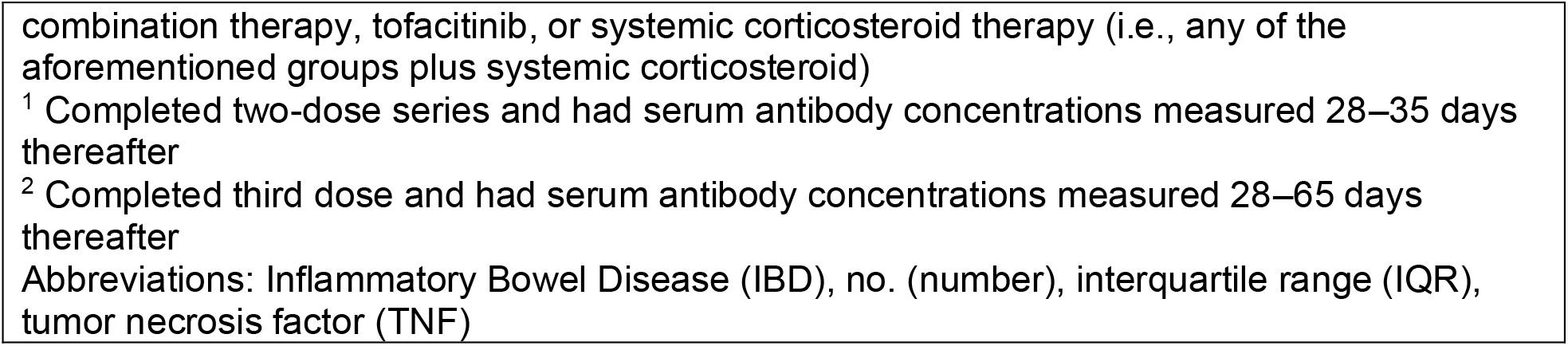
Study participant characteristics

| Table 1: Study participant characteristics |  |  |  |
| --- | --- | --- | --- |
|  | IBD subjects<br>Third dose<br>(n=85)<sup>1</sup> | IBD subjects<br>Two-dose<br>series (n=139)<sup>2</sup> | p-value |
| **Demographics** |  |  |  |
| Age [years] – median (IQR) | 48 (38–60) | 41 (34-52) | 0.003 |
| Male – no. (%) | 38 (44.7) | 71 (51) | 0.35 |
| **Type of IBD** |  |  |  |
| Crohn’s Disease – no. (%) | 55 (64.7) | 96 (69.1) | 0.44 |
| Ulcerative Colitis – no. (%) | 30 (35.3) | 42 (32.2) |  |
| IBD Unclassified | - | 1 (0.7) |  |
| **Vaccine data** |  |  |  |
| Two-dose series |  |  |  |
| Moderna – no. (%) | 37 (43.5) | 67 (48.2) | 0.58 |
| Pfizer – no. (%) | 48 (56.5) | 72 (51.8) |  |
| Third dose |  |  |  |
| Moderna – no. (%) | 35 (41.2) | - |  |
| Pfizer – no. (%) | 50 (58.8) | - |  |
| Time between completion of two-dose series and third dose [days] – median (IQR) | 149 (132–167) | - |  |
| **IBD treatment** |  |  |  |
| Non-systemic immunosuppression<sup>a</sup> – no. (%) | 24 (28.2) | 31 (22.3) | 0.34 |
| Aminosalicylate or no IBD therapy – no. (%) | 3 (3.5) | 17 (12.2) |  |
| Vedolizumab monotherapy – no. (%) | 21 (24.7) | 14 (10.1) |  |
| Duration of therapy [months] – median (IQR)<sup>b</sup> | 36 (17-54) | 26 (16-47) | 0.41 |
| Systemic immunosuppression<sup>c</sup> – no. (%) | 61 (71.8) | 108 (77.7) | 0.34 |
| Thiopurine monotherapy – no. (%) | 6 (7.1) | 14 (10.1) |  |
| Anti-TNF monotherapy – no. (%) | 31 (36.5) | 59 (42.4) |  |
| Anti-TNF combination therapy – no. (%) | 12 (14.1) | 12 (8.6) |  |
| Ustekinumab monotherapy or combination therapy – no. (%) | 9 (10.6) | 15 (10.8) |  |
| Tofacitinib monotherapy – no. (%) | 2 (2.4) | 6 (4.3) |  |
| Systemic corticosteroid therapy – no. (%) | 1 (1.2) | 8 (5.8) |  |
| Duration of therapy [months] – median (IQR) | 42 (18-117) | 43 (14-85) | 0.76 |
| **Serum antibody concentrations** |  |  |  |
| Detectable antibody concentrations – no. (%) | 85 (100) | 135 (97.1) | 0.12 |
| Serum antibody concentrations [mg/mL] – median (IQR) | 68 (32–147) | 31 (16–6.1) | <0.001 |
| Time between vaccine dose and antibody measurement [days] – median (IQR) | 37 (32–47) | 32 (29–34) | <0.001 |
| <sup>a</sup> Absence of IBD therapy, mesalamine monotherapy, or vedolizumab monotherapy |  |  |  |
| <sup>b</sup> Subjects with absence of IBD therapy were omitted from the calculation of duration of therapy |  |  |  |
| <sup>c</sup> Thiopurine monotherapy (i.e., azathioprine, mercaptopurine), anti-TNF monotherapy, anti-TNF combination therapy (i.e., plus antimetabolite), ustekinumab monotherapy or |  |  |  |

combination therapy, tofacitinib, or systemic corticosteroid therapy (i.e., any of the aforementioned groups plus systemic corticosteroid)
<sup>1</sup> Completed two-dose series and had serum antibody concentrations measured 28–35 days thereafter
<sup>2</sup> Completed third dose and had serum antibody concentrations measured 28–65 days thereafter
Abbreviations: Inflammatory Bowel Disease (IBD), no. (number), interquartile range (IQR), tumor necrosis factor (TNF)

In patients with IBD, antibody concentrations were significantly higher following a third dose in comparison to the two-dose series (median 68 (IQR 32–147) vs 31 (IQR 16–61), p<0.001) (Figure 1A). One hundred thirty-five patients with IBD (97.1%) had detectable antibody concentrations at t_1_, while all 85 (100%) had detectable antibody concentrations at t_2_ (p=0.12). Of the two patients with IBD who were seronegative at t_1_ and received a third dose, each had detectable antibody concentrations (mean 6.25 µg/mL, SD 2.1) at t_2_; one subject was on anti-TNF monotherapy and the other was on tofacitinib.

**Figure 1:**
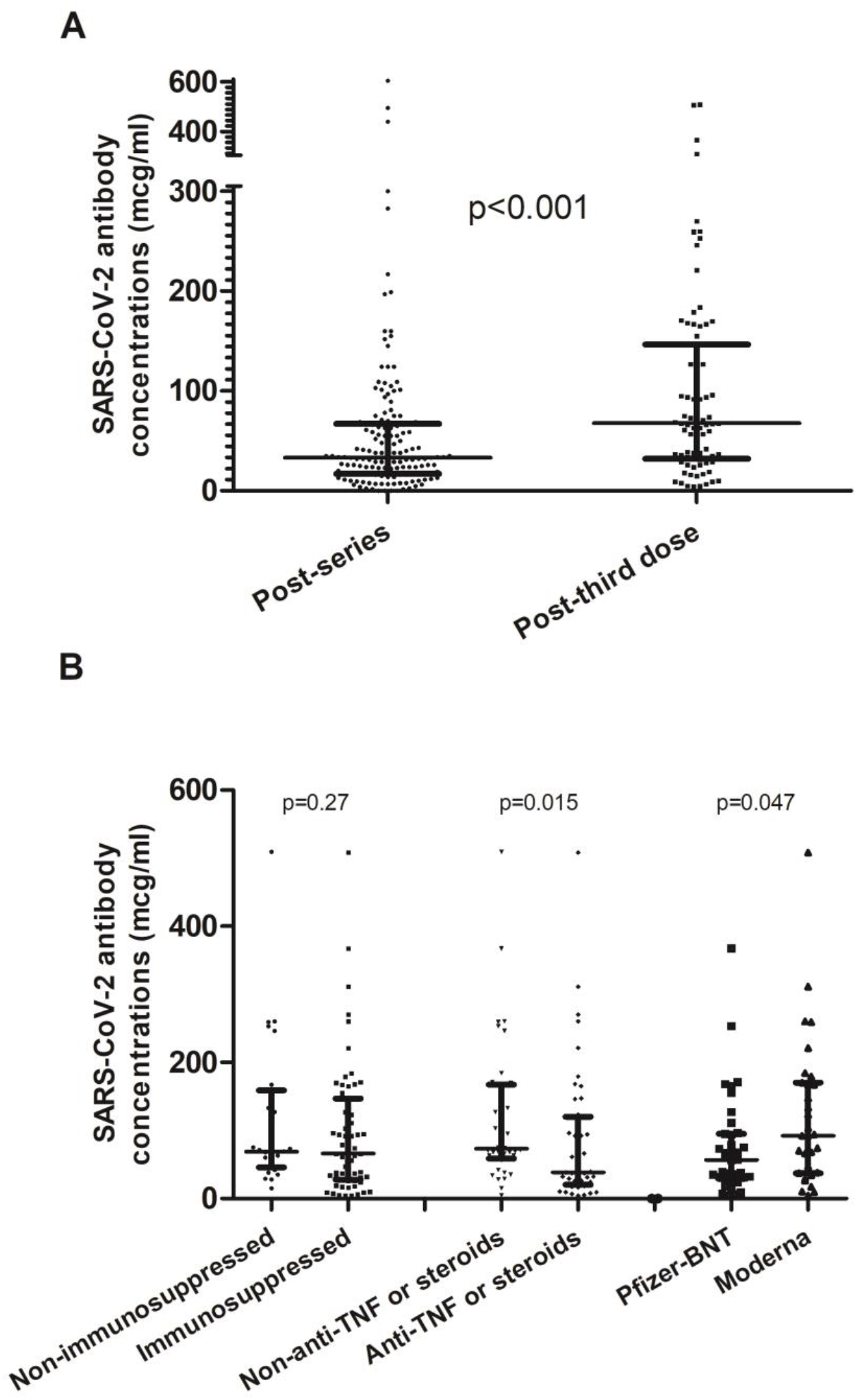
**A**. Serum antibody concentrations in patients with IBD following the two-dose series vs third dose (median 31 (IQR 16–61) vs 68 (IQR 32–147), p<0.001). **B**. Following the third dose in patients with IBD, antibody concentrations in those on immunosuppressive therapy vs non-immunosuppressive therapy (median 69 (IQR 46–159) vs 66 (IQR 28–147), p=0.27); those on anti-TNF monotherapy, anti-TNF combination therapy, and corticosteroid therapy groups vs patients with IBD not on such therapy (median 38 (IQR 20–120) vs 73 (IQR 58–167), p=0.015); and those that received three Moderna doses vs three Pfizer doses (median 94 (IQR 38–170) vs 62 (IQR 31–96), p=0.047). Please see Methods section for definition of medication categories.

At t_2_, antibody concentrations were similar between patients with IBD on immunosuppressive therapy and non-immunosuppressive therapy (median 69 (IQR 46–159) vs 66 (IQR 28–147), p=0.27) (Figure 1B). Further subgroup analysis revealed that those on systemic corticosteroids or anti-TNF combination therapy had significantly lower antibody concentrations at t_2_ than patients that were not (median 29 (IQR 10–39) vs 72 (IQR 37–164), p<0.001). When adding those on anti-TNF monotherapy to the former group, the difference in median antibody concentrations was reduced but remained statistically significant (median 38 (IQR 20–120) vs 73 (IQR 58–167), p=0.015). Serum antibodies were significantly higher at t_2_ for patients with IBD who received three Moderna doses compared to those who received three Pfizer doses (median 94 (IQR 38– 170) vs 62 (IQR 31–96), p=0.047).

Although HC had higher antibody concentrations compared to IBD subjects at t_1_ (median 120 (IQR 88–190) vs 31 (IQR 16–61), p<0.001), HC had lower antibody concentrations than IBD subjects at t_2_ (median 17 (IQR 11–22) vs 68 (IQR 32–147), p<0.001).

## Discussion

All patients with IBD who received a third COVID-19 mRNA vaccine dose were seropositive. Median antibody concentrations were higher following a third dose than after the two-dose series. Patients on corticosteroids and patients on anti-TNF monotherapy or combination therapy had relatively lower antibody concentrations than patients who were not, suggesting that such therapy may blunt the humoral immune response to the vaccine. These findings are similar to those reported in other immunosuppressed patient populations with autoimmune disease, but are distinct from solid organ transplant since all patients seroconverted following three COVID-19 mRNA vaccine doses.^4,5^ Furthermore, those who completed a three-dose Moderna series had higher antibody concentrations than those who completed a three-dose Pfizer series, which is similar to previously reported findings regarding the two-dose series.^2^

There is evidence of waning humoral immunity in the general population following COVID-19 vaccination, especially in those who are immunosuppressed or older than 65 years.^6^ We observed this phenomenon in our HC cohort, none of whom received a third dose. A high incidence of breakthrough infection among vaccinated healthcare workers and the general population has also been described.^7,8^ As such, the ACIP recommended a booster mRNA vaccine dose six months following completion of the original two-dose vaccine series for all adults. Booster doses have reduced incidence of infection and severity of illness.^9,10^ Those with moderate-to-severe immunosuppression who completed a three-dose mRNA vaccine primary series may be eligible for a fourth, booster dose 6 months after the most recent dose. Our data indicate that certain patients with IBD on immune-modifying therapy may benefit from a fourth COVID-19 mRNA vaccine dose. Whether patients with IBD would benefit from a mix-and-match strategy or should continue with their original series when receiving their third or fourth doses of COVID-19 mRNA vaccines is unknown.

Our study has several strengths. We evaluated humoral immunogenicity in patients on stable medication regimens (median 39 months in those who received a third dose), which permitted us to assess the effect of medications on the immune response to vaccination. We also included a HC reference population that received the two-dose primary series but did not receive a third dose. Our study is limited in its sample size, small representation of certain treatment regimens, and the absence of a reference HC population that received a third dose.

In conclusion, all patients with IBD exhibited a humoral immune response following a third COVID-19 mRNA vaccine dose, and this response may be blunted by certain immune-modifying therapy. The role of serum antibody concentrations as a correlate of immunity has not been definitively established. Further studies are needed to investigate the durability of humoral immunity in addition to other aspects of the adaptive immune response following COVID-19 vaccination in the IBD population.

## Data Availability

All data produced in the present work are contained in the manuscript.

## Acknowledgements

The authors thank all the subjects who participated in the study, the specialty pharmacists at UW-Health for their help, and the staff at the Office of Clinical Trials at University of Wisconsin-Madison for all their work.

